# Are allergic diseases a risk factor for systemic side effects after COVID-19 vaccines?

**DOI:** 10.1101/2022.01.23.22269704

**Authors:** Emel Atayik, Gökhan Aytekіn

## Abstract

**Background/ aim:** Mass vaccination seems to be the most effective way to turn back to the pre-pandemic period and end the pandemic. Unfortunately, COVID-19 vaccines have some side effects. In phase studies of currently-approved COVID-19 vaccines, patients with a known allergy or a history of anaphylaxis were excluded from the studies. This situation creates doubts about the course of atopy and the presence of allergic disease related to the side effects of COVID-19 vaccines in patients with allergic diseases. Therefore, our aim with this study was to evaluate local side effects (LSE) and systemic side effects (SSE) after COVID-19 vaccines in patients with allergic diseases and to determine possible risk factors.

**Materials and Methods:** Six hundred forty-eight adult patients who received any COVID-19 vaccine between April 1, 2021 and September 30, 2021 and agreed to participate in the study were included in this case-control retrospective study.

**Results:** Six hundred forty-eight adult patients [Female: 446 (68.8%), Male: 202 (32.2%)] participated in the study. After the 1st dose of COVID-19 vaccine, 24.1% of patients reported SSE. After the 2nd dose of COVID-19 vaccine, 67 patients (12.3%) developed SSE. Female gender (OR: 1.757, 95%Cl: 1.143-2.702, p: 0.010), history of previous COVID-19 infection (OR: 1.762, 95%Cl: 1.068-2.906, p: 0.026), and COVID-19 vaccine type administered (OR: 4.443, 95% CI: 2.640-7.476, p<0.001) were found to be independent risk factors for SSE after COVID-19 vaccines. Premedication (OR: 0.454, 95% Cl: 0.281-0.733, p<0.001), was found to be a protective factor for SSE developing after COVID-19 vaccines.

**Conclusion:** CoronoVac and Pfizer-BioNTech COVID-19 vaccines are shown to be well tolerated. Patients with allergic disease do not have an increased risk for SSE that may develop after COVID-19 vaccines. Moreover, doubts or fears about possible side effects in the allergic patient group should not be an obstacle to COVID-19 vaccination.

## 1. Introduction

Severe acute respiratory syndrome coronavirus 2 (Sars-CoV-2) was first reported in December 2019 and then spread all over the world in a short time and was accepted as a pandemic by the World Health Organization (WHO) in March 2020. In the 2 years since it was first identified, coronavirus disease 2019 (COVID-19) has killed more than 5 million people (1). There is still no effective treatment for the disease. Thus, herd immunity (mass vaccination and herd immunity) for virus protection seems to be the most effective way to turn back to the pre-pandemic period and end the pandemic. However, due to sociodemographic inequalities, vaccine supply problems, vaccine hesitancy; unfortunately, COVID-19 vaccination does not have the desired level and effect.

In Turkey, the first COVID-19 vaccination was started on January 13, 2021, with CoronoVac (Sinovac Biotech, China) to healthcare workers, and then the Pfizer-BioNTech COVID-19 vaccine was added to the vaccination calendar. As of April 2020, individuals aged 60 and over have started to be vaccinated. As of December 2021, the total number of COVID-19 vaccines administered in Turkey is 125 million doses.

Unfortunately, COVID-19 vaccines also have side effects. Although the side effects reported with COVID-19 vaccines are usually minor and can be easily controlled with necessary interventions (2), these side effects lead to doubts about COVID-19 vaccines in patients during the periods when COVID-19 vaccination should be done most rapidly and intensively. Additionally, the rapid development and production phases of COVID-19 vaccines and the limited data of the post-vaccination period increase these doubts.

In phase 2 and 3 of currently-approved COVID-19 vaccines, patients with a known allergy or a history of anaphylaxis were excluded from the study (3). After the first Pfizer-BioNTech COVID-19 vaccine was administered, 2 anaphylaxis cases were reported in the media before 24 hours had passed (4). Also, during December 14–23 2020, 175 severe allergic reactions were reported after approximately 2 million Pfizer-BioNTech COVID-19 vaccine administrations, and 21 of these cases were reported to be anaphylaxis (5). This situation creates doubts and unanswered questions about the course of atopy and the presence of allergic disease related to the side effects of COVID-19 vaccines in patients with allergic diseases as well as clinicians dealing with this patient group (6). There are studies on the efficacy and safety of COVID-19 vaccines, however, studies on the course of COVID-19 vaccines in allergic patients are limited. Therefore, our aim with this study was to evaluate local and systemic side effects after COVID-19 vaccines in patients with allergic diseases, especially allergic rhinitis, asthma, and chronic urticaria, and to determine possible risk factors for these side effects.

## 2. Materials and Methods

Among the patients who applied to the tertiary allergy and immunology clinic between April 1, 2021 and September 30, 2021, 648 adult patients who received any COVID-19 vaccine and agreed to participate in the study were included in this case-control retrospective study.

An anonymous self-reporting survey related to safety and tolerance of vaccine was applied in person to the patients included in the study. This survey is a questionnaire based on patient statements, including demographic data of patients and questions about local side effects (LSE) and systemic side effects (SSE) developed after vaccination. Pain, redness, swelling at the vaccination site after COVID-19 vaccinations were accepted as LSE, and post-vaccine symptoms such as weakness, fatigue, myalgia, arthralgia, fever, headache were accepted as SSE. Age, gender, history of previous COVD-19 infection, COVID-19 vaccine type and number of doses, atopy status of patients who had COVID-19 vaccine, use of a drug that may affect the COVID-19 vaccine side effects, such as antihistamine, steroid, omalizumab, before the COVID-19 vaccine and demographic characteristics such as the presence of a doctor-diagnosed allergic disease were questioned in this survey. The local and systemic side effects developed after vaccination, the development time of the developed side effects, and the need for treatment for the side effects were also questioned. Systemic symptoms such as urticaria-angioedema, shortness of breath, nausea-vomiting, hypotension, syncope and presyncope after COVID-19, which may raise the suspicion of anaphylaxis, and the need for adrenaline were asked. Additionally, patients were asked whether or not they had had a post-COVID-19 vaccine post-COVID-19 infection. Information on atopy status, allergen sensitivity and allergic diseases of the patients were obtained from their files.

Patients who were treated with drugs that have the potential to affect local and systemic side effects such as anti-histamine, oral steroid or omalizumab, and patients who were treated with anti-histamine and/or steroid therapy to avoid the development of side effects before the COVID-19 vaccine were considered as the patient group who received premedication.

Ethics committee approval of the study was granted by Karatay University Ethics Committee (Date: 15.10.2021, decision 2021/012). Written informed consent was obtained from all patients participating in the study.

IBM SPSS 20.0 (Chicago, IL, USA) statistics software was used for the analysis of all data obtained during the study and recorded in the study form. Kolmogorov Smirnov test was used to determine whether or not the distribution of discrete and continuous numerical variables was in accordance with the normal distribution. Descriptive statistics were demonstrated as mean±standard deviation (SD) or median (minimum-maximum) for discrete and continuous numerical variables, and as number of cases and (%) for categorical variables. Chi-square was used to evaluate categorical variables, and t test or Mann Whitney U test was used to evaluate continuous variables. Independent risk factors for LSE and SSE were determined by univariant and multivariant binominal regression analysis. For p<0.05, the results were considered statistically significant.

## 3. Results

Six hundred forty-eight adult patients [Female (F): 446 (68.8%), Male (M): 202 (32.2%)] participated in the study. The median age of the study group was 41 (18-86). 14.8% of the patients (96 patients) have a history of previous COVID-19 infection. Pfizer-BioNTech vaccine was administered to 68.5% of the patients, and double-dose COVID-19 vaccine was administered to 84% (544 patients). Ninety-six patients (14.8% of the patients) were premedicated with anti-allergic drugs before the COVID-19 vaccine. Among the study population, 35.2% were followed up with allergic rhinitis, 22.8% with asthma, and 26.5% with chronic urticaria (Figure 1).

**Figure 1:**
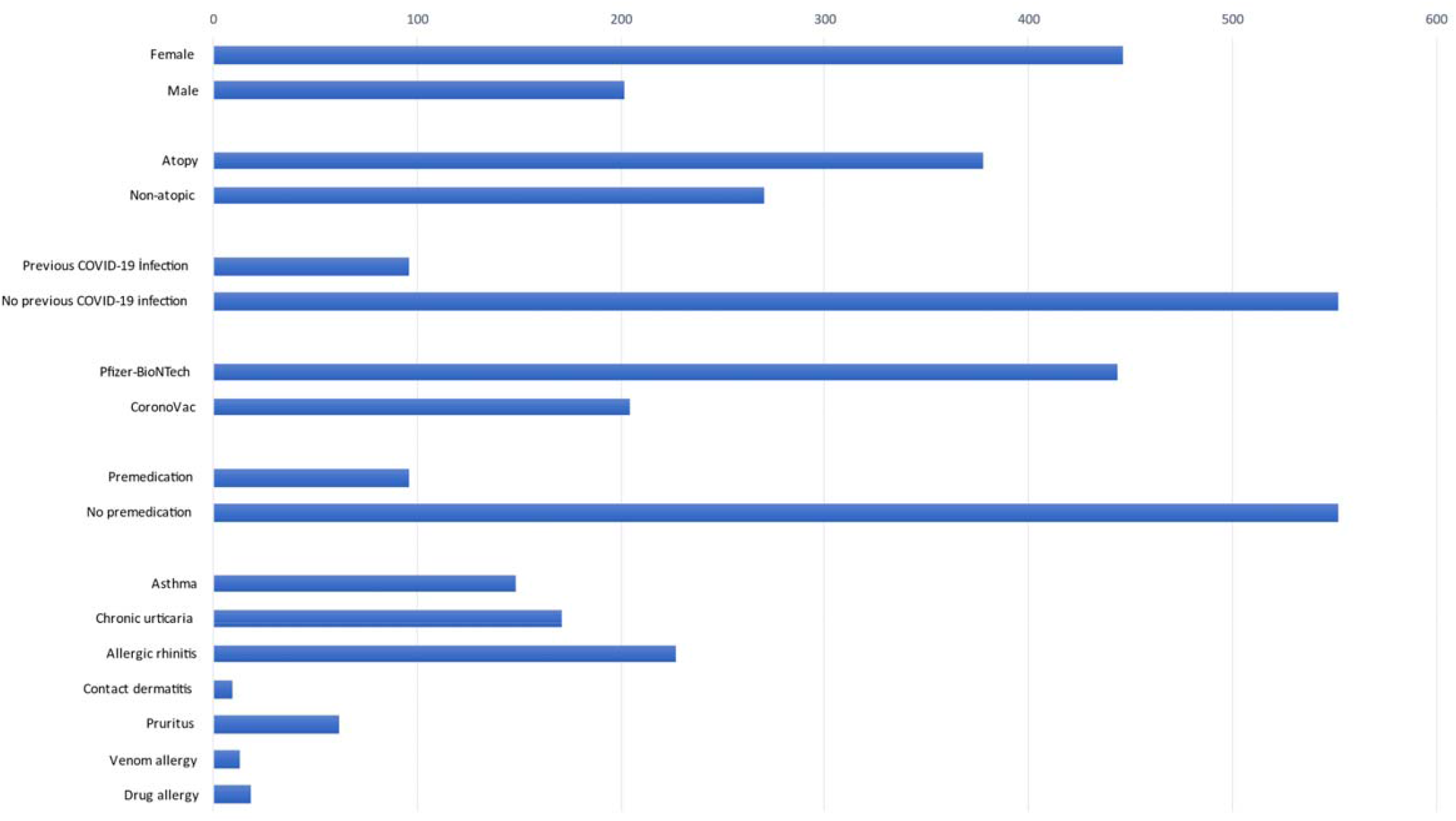
Demographic and clinical parameters of study population COVID-19: Coronavirus disease 2019

Two hundred ninety-three patients (45.2% of the patients) reported that they developed side effects after the 1st dose of COVID-19 vaccine. After the 1st dose of COVID-19 vaccine, 35% of patients had LSE. LSE was most commonly reported in the first 4 hours (26.9%) as local pain at the injection site (34.7%). One hundred fifty-six patients (24.1% of patients) reported SSE after 1st dose of COVID-19 vaccine. SSE occurred most frequently as fatigue (14.5%) within 24-72 hours (13.9%) after the COVID-19 vaccine. Among the patients who reported developing SSE, 44 (6.8%) patients needed treatment with antipyretic and anti-inflammatory drugs. Anaphylaxis developed in two patients (0.3%) after the 1st dose of COVID vaccine and adrenaline injection was administered to these patients.

The number of patients who were administered the 2nd dose of COVID-19 vaccine is 544. After the 2nd dose of COVID-19 vaccine, 104 patients (19.1%) developed LSE, and the most frequently reported LSE was local pain at the injection site (18.9% of patients who received 2nd dose of COVID-19 vaccine - 99% of patients who developed LSE-103 patients). LSE most frequently developed (15.8% of patients who received the 2nd dose of COVID-19 vaccine - 75.4% of patients who developed LSE - 86 patients) after the 4th hour of COVID-19 vaccine injection. Among the 544 patients who were administered the 2nd dose of COVID-19 vaccine, 67 patients (12.3%) developed SSE. SSE occurred most frequently at 24-72 hours after the injection (9.7% of those who received the 2nd dose of COVID-19 vaccine, 79.1% of the patients who reported SSE after the 2nd dose of COVID-19, 53 patients), and it was most commonly reported as fatigue (9.1% of those who received the 2nd dose of COVID-19 vaccine, 73.1% of the patients who reported SSE after the 2nd dose of COVID-19, 49 patients) and arthralgia (7.9% of those who received the 2nd dose of COVID-19 vaccine, 64.2% of the patients who reported SSE after the 2nd dose of COVID-19, 43 patients) (Figure 2). Twelve patients needed treatment with antipyretic and anti-inflammatory drugs after the 2nd dose of COVID-19 vaccine, but no case of anaphylaxis was reported after the 2nd dose of COVID-19 vaccine, and no patient was administered adrenaline injection. Thirteen patients (2%) developed COVID-19 infection despite being vaccinated against COVID-19. The demographic and clinical characteristics of the patients are summarized in Table 1.

**Table 1:** Demographic, clinical and laboratory parameters of the study population and characteristics of LSE and SSE developing after COVID-19 vaccines

| Parameters | Results | After 1st dose of COVID-19 vaccine n: 648 | Results | After 2nd dose of COVID-19 vaccine n: 544 | Results |
| --- | --- | --- | --- | --- | --- |
| Gender, Female, n % | 446 (68.8) | LSE | 227 (35) | LSE | 104 (19.1) |
| Age, year | 41 (18-86) | SSE | 156 (24.1) | SSE | 67 (12.3) |
| Atopy, n % | 378 (58.3) | **Type of LSE, n %** |  | **Type of LSE, n %** |  |
| Previous COVID-19 infection, n % | 96 (14.8) | Redness | 6 (0.9) | Redness | 2 (0.4) |
| COVID-19 vaccines, n % |  | Local pain at injection site | 225 (34.7) | Local pain at injection site | 103 (18.9) |
| Pfizer-BioNTech | 444 (68.5) | Swelling at injection site | 19 (2.9) | Swelling at injection site | 7 (1.3) |
| CoronoVac | 204 (31.5) | **Time of LSE, n %** |  | **Time of LSE, n %** |  |
| COVID-19 vaccine doses, n % |  | Hyper acute (in 30 minutes): | 3 (0.5) | Hyper acute (in 30 minutes): | 2 (1.9) |
| Single dose | 104 (16) | Acute (in 4 hours): | 50 (7.7) | Acute (in 4 hours): | 16 (15.4) |
| Double dose | 544 (84) | Late (After 4 hours): | 174 (26.9) | Late (After 4 hours): | 86 (82.7) |
| Premedications, n % | 96 (14.8) | SSE after 1st dose of COVID-19 vaccine | 156 (24.1) | SSE after 2nd dose of COVID-19 vaccine | 67 (12.3) |
| Diagnosis, n % |  | **Type of SSE, n %** |  | **Type of SSE, n %** |  |
| Asthma | 148 (22.8) | Fatigue | 94 (14.5) | Fatigue | 49 (9.1) |
| Chronic urticaria | 171 (26.5) | Arthralgia | 62 (9.6) | Arthralgia | 43 (7.9) |
| Allergic rhinitis | 227 (35.2) | Myalgia | 51 (7.9) | Myalgia | 33 (6.1) |
| Contact dermatitis | 9 (1.4) | Headache | 60 (9.3) | Headache | 28 (5.1) |
| Pruritus | 62 (9.6) | Fever | 45 (6.9) | Fever | 21 (3.9) |
| Venom allergy | 13 (2.0) | 38 °C> | 39 (6) | 38 °C> | 17 (3.2) |
| Drug allergy | 18 (2.8) | 38 °C < | 6 (0.9) | 38 °C < | 4 (0.7) |
|  |  | Urticaria/ Angioedema | 11 (1.7) | Urticaria/ Angioedema | 5 (0.9) |
| COVID-19 infection after vaccination | 13 (2) | Dyspnea/ Wheezing | 12 (1.9) | Dyspnea/ Wheezing | 2 (0.4) |
| After 2 dose of Pfizer-BioNTech | 6 | Nausea/ Vomiting | 14 (2.2) | Nausea/ Vomiting | 4 (0.7) |
| After 2 dose of CoronoVac | 6 | Hypotension/ tachcardia/ syncope/ presyncope | 13 (2) | Hypotension/ tachcardia/ syncope/ presyncope | 3 (0.6) |
| After single dose of BioNTech | 1 | **Time of SSE, n %** |  | **Time of SSE, n %** |  |
|  |  | In 24 hours | 63 (9.7) | In 24 hours | 13 (2.4) |
| Anaphylaxis |  | 24-72 hours | 90 (13.9) | 24-72 hours | 53 (9.7) |
| After 1. dose | 2 (0.3) | 72h-7 days | 2 (0.3) | 72h-7 days | 1 (0.2) |
| After 2. dose | - | After 7 days | 1 (0.2) | After 7 days | - |
|  |  | Treatment | 44 (6.8) | Treatment | 12 (2.2) |

LSE: local side effects, SSE: systemic side effects, COVID-19: Coronavirus disease 2019

**Figure 2:**
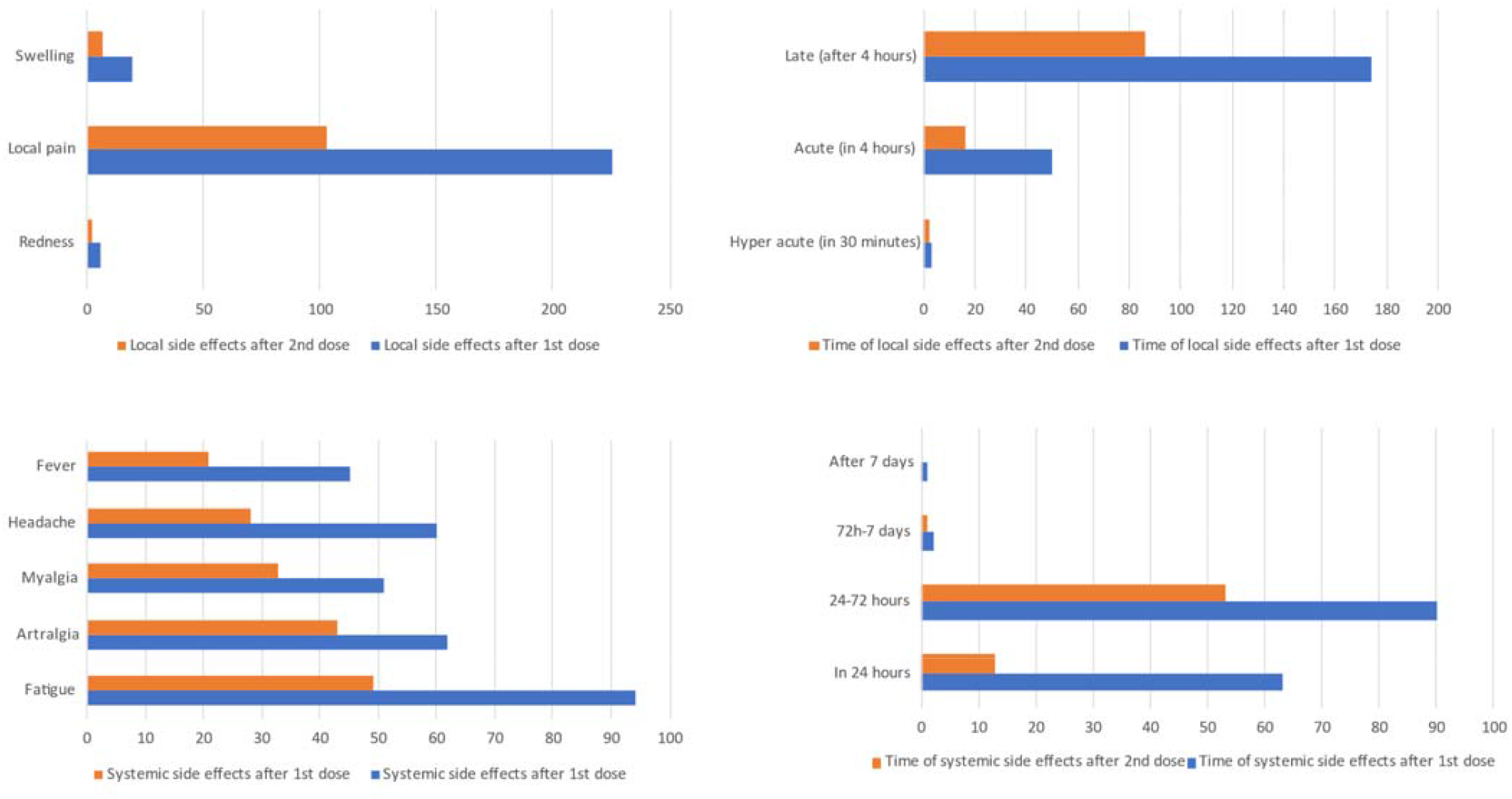
Properties of LSE and SSE after first and second dose of COVID-19 Vaccine LSE: Local side effects, SSE: Systemic side effects, COVID-19: Coronavirus disease 2019

In the comparison of patient groups reporting LSE and not reporting LSE after the first dose of COVID-19 vaccine, a significant difference was found between the two groups in terms of age, the proportion of patients < 50 years of age, the applied COVID-19 vaccines (Coronovac vaccine vs Pfizer-BioNTech vaccine), the rate of premedicated patients, and the presence of allergic rhinitis (p<0.001, p<0.001, p<0.001, p<0.001, p<0.001 and p: 0.002), respectively. On the other hand, there was a significant difference between patients who reported SSE after the first dose of COVID-19 vaccine (Coronovac vaccine vs Pfizer-BioNTech vaccine) and those who did not, in terms of gender, the proportion of patients < 50 years old, history of previous COVID-19 infection, applied COVID-19 vaccines, and premedication status (respectively p: 0.021, p: 0.012, p: 0.021, p<0.001, p<0.001, p< 0.001) (Table 2) (Figure 3).

**Table 2:** Comparison of LSE and SSE reported after COVID-19 vaccines with demographic and clinical characteristics of patients

|  | After 1st dose of COVID-19 vaccines |  |  |  |  |  | After 2nd dose of COVID-19 vaccines |  |  |  |  |  |
| --- | --- | --- | --- | --- | --- | --- | --- | --- | --- | --- | --- | --- |
|  | LSE (+)<br>n: 227 | LSE (-)<br>n: 421 | p | SSE (+)<br>n: 156 | SSE (-)<br>n: 492 | p | LSE (+)<br>n: 104 | LSE (-)<br>n: 440 | p | SSE (+)<br>n: 67 | SSE (-)<br>n: 477 | p |
| **Gender, female, n (%)** | 167 (73.6) | 279 (66.3) | 0.056 | 119 (76.3) | 327 (66.5) | **0.021** | 74 (71.2) | 294 (66.8) | 0.395 | 51 (76.1) | 317 (66.5) | 0.113 |
| **Age, year** | 38 (17-71) | 43 (17-86) | **<0.001** | 40.50 (18-70) | 41 (18-86) | 0.463 | 40.5 (17-71) | 42 (17-86) | 0.264 | 40 (18-70) | 42 (17-86) | 0.487 |
| **Age (<50 year)** | 181 (79.7) | 260 (61.8) | **<0.001** | 119 (76.3) | 322 (65.4) | **0.012** | 78 (75.0) | 281 (63.9) | **0.031** | 52 (77.6) | 307 (64.4) | **0.032** |
| **Presence of atopy, n (%)** | 146 (64.3) | 232 (55.1) | 0.230 | 101 (64.7) | 277 (56.3) | 0.062 | 67 (64.4) | 251 (57.0) | 0.170 | 43 (64.2) | 275 (57.7) | 0.310 |
| **History of previous COVID-19 infection, n (%)** | 40 (17.6) | 56 (13.3) | 0.140 | 32 (20.5) | 64 (13.0) | **0.021** | 20 (19.2) | 56 (12.7) | 0.085 | 23 (34.2) | 53 (11.1) | **<0.001** |
| **Coronovac vaccine, n (%)** | 25 (11.0) | 179 (42.5) | **<0.001** | 19 (12.2) | 185 (37.6) | **<0.001** | 12 (11.5) | 170 (38.6) | **<0.001** | 8 (11.9) | 174 (36.5) | **<0.001** |
| **Pfizer-BioNTech vaccine, n (%)** | 202 (89) | 242 (57.5) | **<0.001** | 137 (87.8) | 307 (62.4) | **<0.001** | 92 (88.5) | 270 (61.4) | **<0.001** | 59 (88.1) | 303 (63.5) | **<0.001** |
| **Premedication, n (%) °** | 55 (24.2) | 41 (9.7) | **<0.001** | 38 (24.4) | 58 (11.8) | **<0.001** | 31 (29.8) | 24 (5.5) | **<0.001** | 17 (25.4) | 38 (8.0) | **<0.001** |
| **Asthma, n (%)** | 42 (18.5) | 106 (25.2) | 0.053 | 37 (23.7) | 111 (22.6) | 0.764 | 19 (18.3) | 114 (25.9) | 0.103 | 16 (23.9) | 117 (24.5) | 0.908 |
| **Urticaria, n (%)** | 52 (22.9) | 120 (28.5) | 0.124 | 43 (27.6) | 129 (26.2) | 0.740 | 31 (29.8) | 127 (28.9) | 0.849 | 18 (26.9) | 140 (29.4) | 0.675 |
| **Rhinitis, n (%)** | 98 (43.2) | 130 (30.9) | **0.002** | 56 (35.9) | 172 (35.0) | 0.831 | 46 (44.2) | 133 (30.2) | **0.006** | 25 (37.3) | 154 (32.3) | 0.412 |
| **Pruritus, n (%)** | 27 (11.9) | 35 (8.3) | 0.139 | 16 (10.3) | 46 (9.3) | 0.737 | 6 (5.8) | 34 (7.7) | 0.491 | 7 (10.4) | 33 (6.9) | 0.300 |
| **Drug allergy, n (%)** | 3 (1.3) | 15 (3.6) | 0.132 | 2 (1.3) | 16 (3.3) | 0.267 | 2 (1.9) | 16 (3.6) | 0.380 | 2 (3.0) | 16 (3.4) | 0.874 |
| **Venom allergy, n (%)** | 4 (1.8) | 9 (2.1) | 0.745 | 3 (1.9) | 10 (2.0) | 0.932 | 1 (1.0) | 9 (2.0) | 0.459 | 1 (1.5) | 9 (1.9) | 0.822 |
| **Contact dermatitis, n (%)** | 3 (1.3) | 6 (1.4) | 0.914 | 1 (0.6) | 8 (1.6) | 0.360 | 1 (1.0) | 7 (1.6) | 0.632 | 0 | 8 (1.7) | 0.604 |

LSE: local side effects, SSE: systemic side effects, COVID-19: Coronavirus disease 2019, °Premedication before vaccination

**Figure 3:**
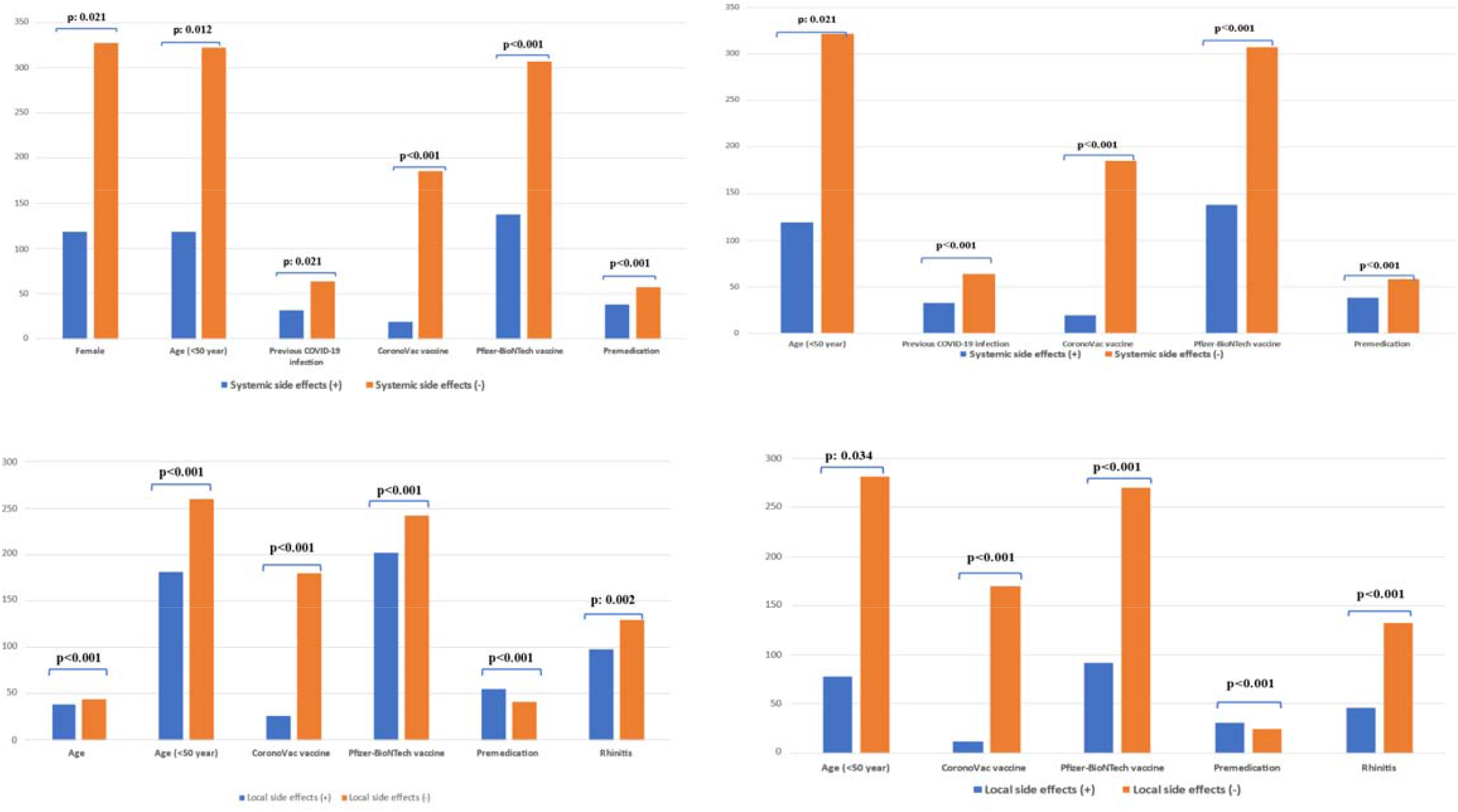
Comparison of LSE and SSE reported after COVID-19 vaccines with demographic and clinical characteristics of patients LSE: Local side effects, SSE: Systemic side effects, COVID-19: Coronavirus disease 2019

In the comparison of the patient groups that did not develop LSE and LSE after the 2nd dose of covid vaccine, a significant difference was found between the two groups in terms of the rate of patients < 50 years of age, the administered COVID-19 vaccines (Coronovac vaccine vs Pfizer-BioNTech vaccine), the rate of premedicated patients, and the presence of allergic rhinitis (respectively p: 0.031, p< 0.001, p< 0.001, p< 0.001, p: 0.006). On the other hand, a significant difference was found between patients who reported SSE after the second dose of COVID-19 vaccine and those who did not, in terms of the ratio of patients < 50 years of age, history of previous COVID-19 infection, the number of administered COVID-19 vaccines, the rate of premedicated patients (respectively p: 0.032, p< 0.001, p< 0.001, p< 0.001, and p< 0.001) (Table 2) (Figure 3).

As a result of the univariate and multivariate analysis, female gender (female vs male, Odds ratio (OR): 1.757, 95%Cl: 1.143-2.702, p: 0.010), history of previous COVID-19 infection (yes vs no, OR: 1.762, 95%Cl: 1.068-2.906, p: 0.026), and COVID-19 vaccine type administered (Pfizer-BioNTech vaccine vs Coronovac vaccine, OR: 4.443, 95% CI: 2.640-7.476, p<0.001) were found to be independent risk factors for SSE after the 1st dose of COVID-19 vaccine. Premedication (yes vs no, OR: 0.454, 95% Cl: 0.281-0.733, p<0.001), on the other hand, was found to be a protective factor for SSE developing after 1st dose of COVID-19 vaccines (Table 3) (Figure 4).

**Table 3:**
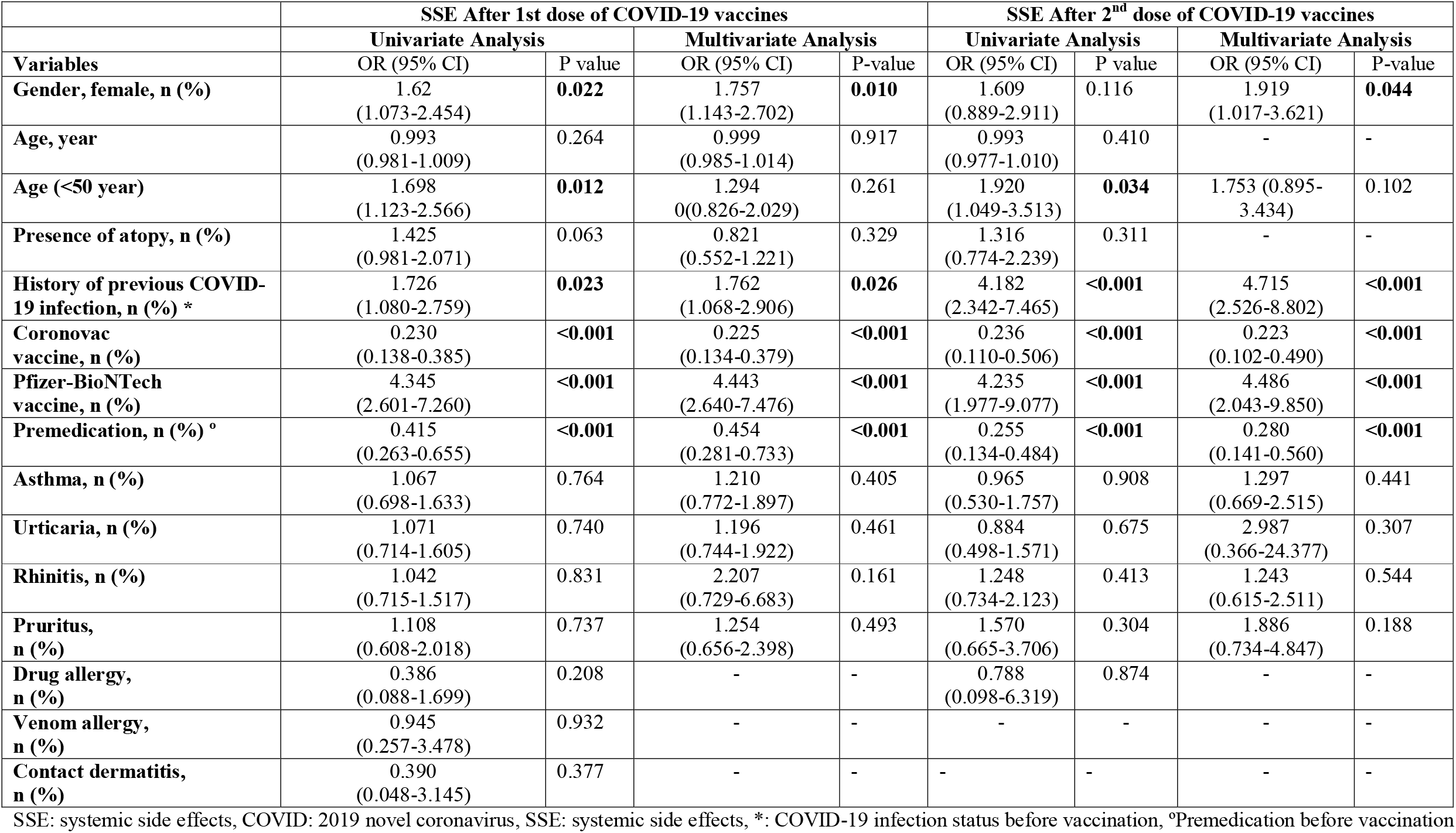
Univariate and multivariate binomial regression analyses demonstrating the relationship between baseline characteristics and SSEs after COVID-19 vaccines

|  | SSE After 1st dose of COVID-19 vaccines |  |  |  | SSE After 2<sup>nd</sup> dose of COVID-19 vaccines |  |  |  |
| --- | --- | --- | --- | --- | --- | --- | --- | --- |
|  | Univariate Analysis |  | Multivariate Analysis |  | Univariate Analysis |  | Multivariate Analysis |  |
| Variables | OR (95% CI) | P value | OR (95% CI) | P-value | OR (95% CI) | P value | OR (95% CI) | P-value |
| Gender, female, n (%) | 1.62<br>(1.073-2.454) | **0.022** | 1.757<br>(1.143-2.702) | **0.010** | 1.609<br>(0.889-2.911) | 0.116 | 1.919<br>(1.017-3.621) | **0.044** |
| Age, year | 0.993<br>(0.981-1.009) | 0.264 | 0.999<br>(0.985-1.014) | 0.917 | 0.993<br>(0.977-1.010) | 0.410 | - | - |
| Age (<50 year) | 1.698<br>(1.123-2.566) | **0.012** | 1.294<br>(0.826-2.029) | 0.261 | 1.920<br>(1.049-3.513) | **0.034** | 1.753 (0.895-3.434) | 0.102 |
| Presence of atopy, n (%) | 1.425<br>(0.981-2.071) | 0.063 | 0.821<br>(0.552-1.221) | 0.329 | 1.316<br>(0.774-2.239) | 0.311 | - | - |
| History of previous COVID-19 infection, n (%) * | 1.726<br>(1.080-2.759) | **0.023** | 1.762<br>(1.068-2.906) | **0.026** | 4.182<br>(2.342-7.465) | **<0.001** | 4.715<br>(2.526-8.802) | **<0.001** |
| Coronovac vaccine, n (%) | 0.230<br>(0.138-0.385) | **<0.001** | 0.225<br>(0.134-0.379) | **<0.001** | 0.236<br>(0.110-0.506) | **<0.001** | 0.223<br>(0.102-0.490) | **<0.001** |
| Pfizer-BioNTech vaccine, n (%) | 4.345<br>(2.601-7.260) | **<0.001** | 4.443<br>(2.640-7.476) | **<0.001** | 4.235<br>(1.977-9.077) | **<0.001** | 4.486<br>(2.043-9.850) | **<0.001** |
| Premedication, n (%) ° | 0.415<br>(0.263-0.655) | **<0.001** | 0.454<br>(0.281-0.733) | **<0.001** | 0.255<br>(0.134-0.484) | **<0.001** | 0.280<br>(0.141-0.560) | **<0.001** |
| Asthma, n (%) | 1.067<br>(0.698-1.633) | 0.764 | 1.210<br>(0.772-1.897) | 0.405 | 0.965<br>(0.530-1.757) | 0.908 | 1.297<br>(0.669-2.515) | 0.441 |
| Urticaria, n (%) | 1.071<br>(0.714-1.605) | 0.740 | 1.196<br>(0.744-1.922) | 0.461 | 0.884<br>(0.498-1.571) | 0.675 | 2.987<br>(0.366-24.377) | 0.307 |
| Rhinitis, n (%) | 1.042<br>(0.715-1.517) | 0.831 | 2.207<br>(0.729-6.683) | 0.161 | 1.248<br>(0.734-2.123) | 0.413 | 1.243<br>(0.615-2.511) | 0.544 |
| Pruritus, n (%) | 1.108<br>(0.608-2.018) | 0.737 | 1.254<br>(0.656-2.398) | 0.493 | 1.570<br>(0.665-3.706) | 0.304 | 1.886<br>(0.734-4.847) | 0.188 |
| Drug allergy, n (%) | 0.386<br>(0.088-1.699) | 0.208 | - | - | 0.788<br>(0.098-6.319) | 0.874 | - | - |
| Venom allergy, n (%) | 0.945<br>(0.257-3.478) | 0.932 | - | - | - | - | - | - |
| Contact dermatitis, n (%) | 0.390<br>(0.048-3.145) | 0.377 | - | - | - | - | - | - |

SSE: systemic side effects, COVID: 2019 novel coronavirus, SSE: systemic side effects, \*: COVID-19 infection status before vaccination, °Premedication before vaccination

**Figure 4:**
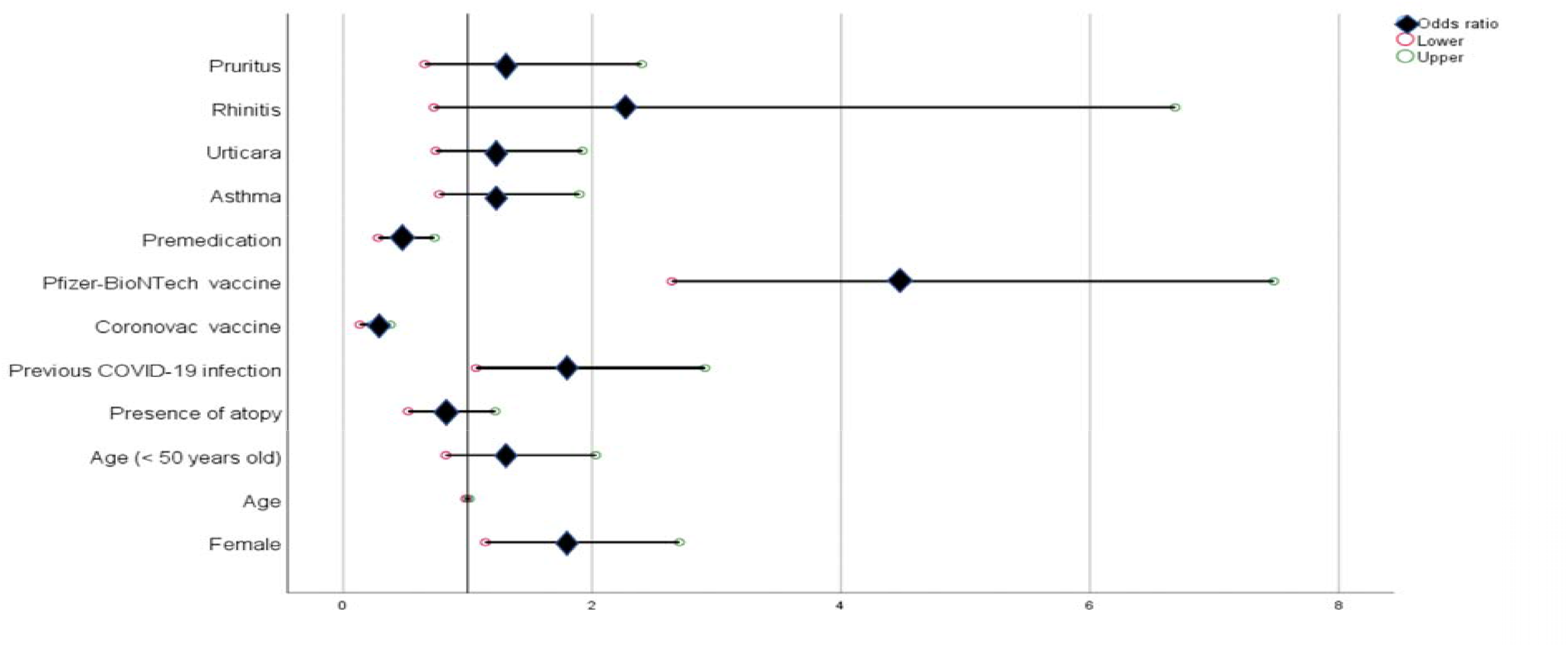
Independent predictors for SSE after first dose of COVID-19 vaccine SSE: Systemic side effects, COVID-19: Coronavirus disease 2019

As a result of univariate and multivariate analysis, female gender gender (female vs male, Odds ratio (OR): 1.919, 95%Cl: 1.017-3.621, p: 0.044), history of previous COVID-19 infection (yes vs no, OR: 4.715, 95%Cl: 2.526-8.802, p<0.001), and the type of COVID-19 vaccine administered (Pfizer-BioNTech vaccine vs Coronovac vaccine, OR: 4.486, 95% CI: 2.043-9.850) were found to be independent risk factors for SSE after the 2nd dose of COVID-19 vaccine. Premedication (yes vs no, OR: 0.280, 95% CI: 0.141-0.560), on the other hand, was found to be a protective factor for SSE developing after 1st dose of CoVid-19 vaccines (Table 3) (Figure 5).

**Figure 5:**
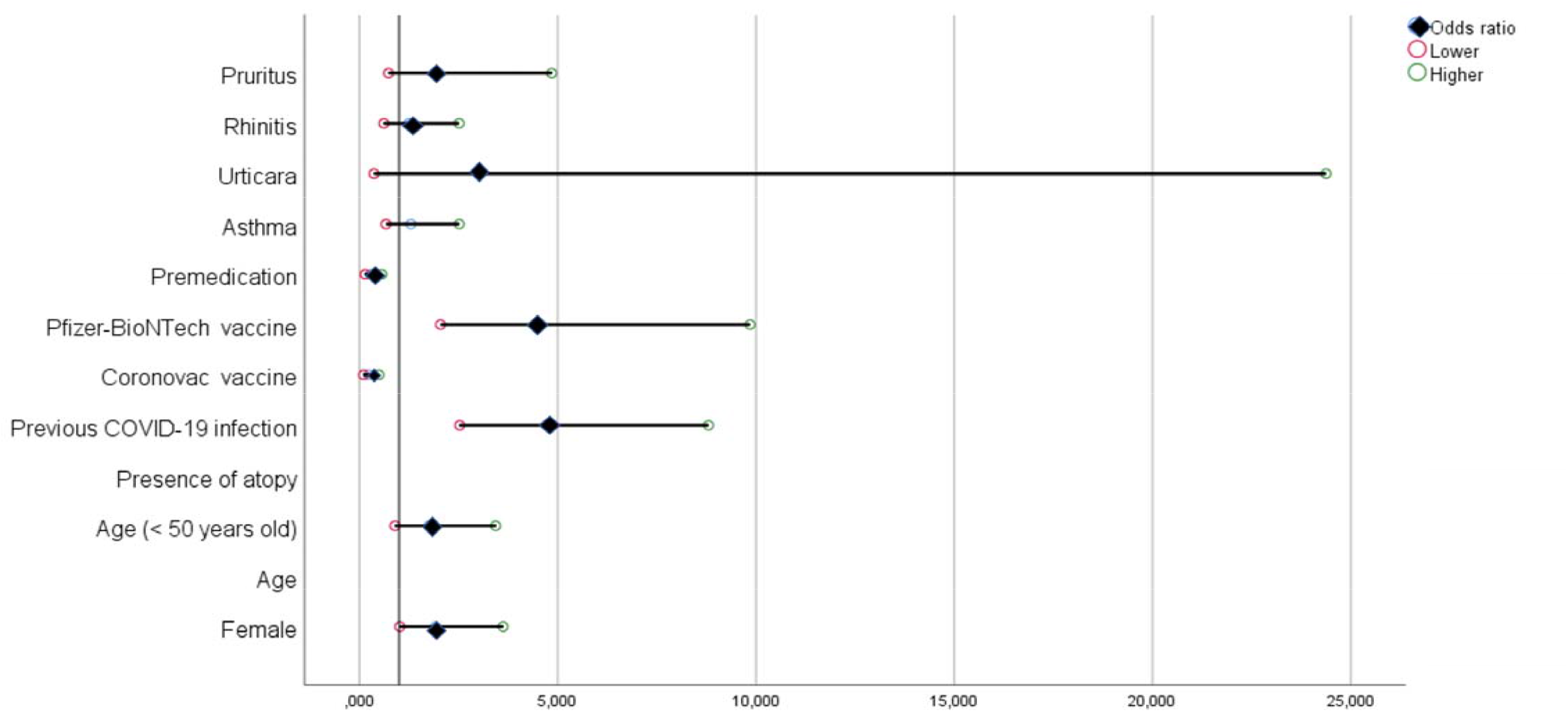
Independent predictors for SSE after second dose of COVID-19 vaccine LSE: Local side effects, SSE: Systemic side effects, COVID-19: Coronavirus disease 2019

## 4. Discussion

To our best knowledge, our study is among the rare studies evaluating the side effects of allergic diseases due to COVID-19 vaccines. The study has 3 main and important results. Firstly, the most common LSE reported after COVID-19 vaccines is injection site pain, and the most common SSE reported is fatigue. Secondly, female gender, history of previous COVID-19 infection, Pfizer-BioNTech vaccine were found to be risk factors for SSE and premedication was found to be protective factors for SSE. Lastly, the presence of allergic disease and atopy, especially allergic rhinitis, asthma and chronic urticaria, is not a risk factor for SSE developing after COVID-19 vaccine.

It has been reported in many studies examining the side effects developed after COVID-19 vaccines that the most common LSE caused by COVID-19 is pain at the injection site, and the most common SSE is weakness/fatigue (7-11). Pain at the injection site was reported as the most common adverse event in the CoronaVac, phase 1 and 2 studies (12). The most frequently reported side effect after another inactive COVID-19 vaccine (BBV152) is pain at the injection site (13). Menni et al. reported the incidence of LSE as 71.9% after 1st dose of Pfizer-BioNTech vaccine and 68.5% after 2nd dose; and the incidence of SSE as 13.5% after the first dose and 22.0% after the second dose (14). Thomas et al reported that the most common LSE after Pfizer-BioNTech vaccine was mild-moderate pain at the injection site, and that fatigue was the most common SSE (15). In another cohort, side effects were reported in 64.9% of 8682 patients who received the first dose of Pfizer-BioNTech or Moderna vaccine, and 80.3% of patients who received the second dose of Pfizer-BioNTech or Moderna vaccine. In this cohort, the most common side effects after COVID-19 vaccines were fatigue and pain at the injection site (16). The findings obtained in our study are also similar to these studies, are compatible with the literature and support the literature.

Anaphylaxis is a common life-threatening allergic reaction. Anaphylaxis after vaccinations is rare and typically emerges within minutes of vaccination (17). The CDC COVID-19 Response Team reported the rate of anaphylaxis after Pfizer-BioNTech vaccine as 11.1 per million (18). This rate is approximately 8 times the risk of anaphylaxis developing due to commonly used vaccines (1.31 per million) (19). Moreover, the rate of anaphylaxis after the first dose of COVID-19 vaccine was found to be 0.3% in our study.

Women generally have stronger immune functions and higher antibody levels, more functional antibodies, but also develop more frequent side effects to vaccines (20). This effect applies to common vaccines such as influenza, yellow fever, rubella, measles, mumps, hepatitis A and B as well as COVID-19 vaccines (21). Many studies also stated that LSE and SSE were reported more frequently in women after COVID-19 vaccines. Lee et al. reported that side effects were more common in women after the 2nd dose of COVID-19 vaccine in healthcare workers who received two doses of Pfzer-BioNTech vaccine in Korea, but a similar relationship was not observed after the first dose of COVID-19 vaccine (8). Menni et al. stated that women reported more side effects after COVID-19 vaccines. (14). In their meta-analysis, Alhumaid et al. found female gender as a risk factor for anaphylaxis and nonanaphylactic reactions (22). In a cohort evaluating side effects related to Pfizer-BioNTech and Moderna vaccines, it was reported that female gender was associated with higher odds in terms of both side effects and severe adverse effects (16). In our study, also, the rate of SSE after both 1st and 2nd dose COVID-19 vaccines was higher in women, but this rate is significant only for SSE developing after the first dose of COVID-19 vaccine. Additionally, the risk of developing SSE in women after COVID-19 vaccines is 1.9 times higher than in men.

The difference in terms of SSE between genders may be caused by genetic, hormonal and immunological differences or the combination of these differences. Women produce a stronger post-vaccine immune response due to innate immunity, increased Toll like receptor-7-mediated recognition, and increased type 1 interferon. Additionally, women are more active in humoral immunity than men. Women showed higher B-cell growth and neutralizing antibody production after vaccinations. Moreover, the effect of sex hormones on gene expression on the X and Y chromosomes influences the immune response to vaccines. Chromosomal mosaicism resulting from random X-chromosome inactivation affects female immune response (23). In addition, it has been claimed that one of the factors affecting the immune response to vaccines or natural infections is the bicrobiome, and that both sex hormones and differences such as diet and lifestyle affect vaccine responses by creating microbiome differences (24)

Previous COVID-19 infection is an important parameter that affects post-COVID-19 vaccine side effects, and in many studies, this relationship has been investigated. Bandolli et al. reported that the rates of LSE after Pfizer-BioNTech vaccine were similar in patients with a history of previous COVID-19 infection compared to patients who did not have COVID-19, but that SSI developed more frequently (25). It was found by Mathioudakis et al. that previous COVID-19 infection is a risk factor for side effects, fever, breathlessness, flu-like illness and fatigue after COVID-19 vaccines (26). Another study stated that previous COVID-19 infection increased the risk of SSE by 2.9 times after Pfizer-BioNTech and a similar relationship was found in LSE (14). Beatty et al. reported the history of previous COVID-19 infection was associated with higher odds (OR, 2.17; 95% CI, 1.77-2.66) (16). They suggested that the reason for this situation is that vaccines increase immunogenicity in infected individuals, COVID-19 vaccines create stronger humoral and T-cell response due to high antibody titers in those with COVID-19 infection and increase post vaccine reactogenity after COVID-19 vaccines in those who have COVID-19 infection (27, 28). Although we did not reach a conclusion in this direction in our study, a clinical observation to support these hypotheses may be to report more frequent side effects after the 2nd dose of COVID-19 vaccines. There are many studies in the literature reporting that more frequent side effects are observed after the second dose of COVID-19 vaccines (8, 14, 16, 26, 29). Sensitized T cells and neutralizing antibodies formed after the first dose of COVID-19 vaccines, may cause a stronger immune response and a higher rate of side effects after a 2nd dose of COVID-19 vaccine – similar to patients who have had a previous COVID-19 infection – which may then explain why COVID-19 infections increase side effects.

CoronoVac is an inactivated COVID-19 vaccine and uses alum with a well-documented safety profile as an adjuvant, which is also used in many other vaccines. Alum increases the neutralizing antibody response, but does not sufficiently stimulate T cells. Therefore, although inactive vaccines are safer in terms of side effects, their effectiveness is shorter and booster doses are needed to maintain their effectiveness. Pfizer-BioNTech COVID-19 vaccine is an mRNA-lipid nanoparticle-based vaccine. It contains polyethylene glycol (PEG) derivatives as an adjuvant, affects both humoral and cellular steps of the adaptive immune system, and its incidence of side effects is relatively higher. Additionally, side effects can be triggered by excipients such as polyethylene glycol (PEG), polysorbates, tromethamine/trometamol used in COVID-19 vaccines. (30-32). In many studies, it has been reported that less LSE and SSE develop with inactive COVID-19 vaccines than with protein subunit vaccines, RNA based vaccines and viral vector vaccines (7, 9, 12). In their meta-analysis, Wu et al. reported that both local and systemic side effects are rarer in inactive COVID-19 vaccines compared to mRNA vaccines (23.7%-89.4% for LSE, 21%-83.3% for SSE) (7). In another meta-analysis, 31.75% of side effects were reported in inactive virus vaccines and 81.76% in RNA-based COVID-19 vaccines (9). Likewise, in our study, both LSE and SSE were observed more frequently after Pfizer-BioNTech COVID-19 vaccine, and Pfizer-BioNTech vaccine was found to be a risk factor for SSE.

Treatments such as anti-histamines, steroid treatments and omalizumab are the most frequently used treatment agents in allergic patients. Premedication with steroids and antihistamines was suggested by Greenhawt et al., albeit at a low level of evidence, as it could theoretically suppress the immune response and reduce side effects after COVID-19 (3). Additionally, it is recommended to consider premedication before COVID-19 vaccines in patients with mastocytosis (33). It was reported by Rama et al. that antihistamine and montelukast premedication and Pfizer-BioNTech vaccine were administered to two healthcare workers with cutaneous and systemic mastocytosis without any side effects (34). Ge et al., on the other hand, suggested that antihistamines protect against post-COVID-19 side effects, as well as that antihistamines have anti-viral effects, and that anti-histamine drugs can bind to ACE2 and prevent the entry of Sars-CoV-2 virus into the cell (35). Similarly, Vila-Corcoles et al. found that antihistamines were protective for COVID-19 in their study on 79083 patients in Spain (36). In another study, it was reported that 70 patients out of 80 who reported allergic complaints after the first dose of Pfizer-BioNTech vaccine received the second dose of vaccine after allergic evaluation, and 89% of the patients developed no reaction or mildly reaction after anti-histamine premedication (37). Unlike this, in a study in Turkey conducted on healthcare workers who received CoronoVac, post-vaccine rash, fever, chills, and headache were reported more frequently in patients receiving anti-histamine (38). Hence, the potential relationship between antihistamine drugs and drug side effects needs further research.

As a result of our study, another finding is that allergic diseases, especially asthma, allergic rhinitis and chronic urticaria, or the presence of atopy are not risk factors for SSE developing after COVID-19 vaccines. Studies on the course of side effects of allergic diseases after COVID-19 vaccines are very limited. In a meta-analysis, Alhumaid et al. found history of atopy anaphylactic and nonanaphylactic reactions to SARS-CoV-2 vaccines (Pfizer-BioNTech and Moderna vaccines) as a risk factor. Inoue et al. reported that although no cases of anaphylaxis were reported, the frequency of adverse events after COVID-19 was higher in allergic patients and the duration of adverse events was longer (39). Nittner et al. reported that local side effects such as swelling and redness develop more frequently in allergic patients after Pfizer-BioNTech vaccine, that the developing side effects last longer, and that allergic individuals need medical intervention 2 times more often than non-allergic individuals due to the side effects (29). On the other hand, Beatty et al. reported that there was asthma associated with lower odds in terms of both side effects and severe adverse effects after Pfizer-BioNTech and Moderna vaccines (16). EACCI (European Academy of Allergy and Clinical Immunology) declared that there is no contraindication for allergic patients to be vaccinated for COVID-19, except for patients with sensitivity to components in COVID-19 vaccines, and that allergy to drugs, food, insect venom, or inhalant allergens (house dust mites, pollens, animal dander, molds) generally does not constitute a contraindication for any vaccine, including SARS-CoV-2 vaccines (40). Moreover, the American College of Allergy Asthma and Immunology (ACAAI) has stated that “Individuals with commune allergies (foods, inhalants, latex, insects) are no more likely than the general public to have an allergic reaction to the Pfizer-BioNTech vaccine” (41). We found in our study that neither the presence of atopy nor the presence of allergic disease was a risk factor for SSE after COVID-19 vaccines, in line with these recommendations.

The strength of our study is that the allergic diseases of the patients were diagnosed by a physician, self-reported allergy is typically much higher than confirmed allergy and the study population size in terms of an error group consisting of allergic patients. Our study also has some limitations. First of all, the reported LSE and SSE were based on the patient’s statement. Threshold values for side effects may vary among patients. Some patients even consider minor side effects, but some patients may not need to report side effects unless they cause very serious complaints. This situation may have been over-or underestimated. Additionally, the number of patients diagnosed with venom allergy, drug allergy and contact dermatitis is relatively low. This limits the power of statistical analyzes in these patient groups.

As a result, in our study, CoronoVac and Pfizer-BioNTech COVID-19 vaccines are shown to be well tolerated. Patients with allergic disease do not have an increased risk for SSE that may develop after COVID-19 vaccines. Clinicians dealing with this patient group should have no doubts in this regard. Moreover, doubts or fears about possible side effects in the allergic patient group should not be an obstacle to COVID-19 vaccination.

## Data Availability

All data produced in the present study are available upon reasonable request to the authors

## Acknowledgement

None

